# Rheumatoid arthritis susceptibility genes show pathotype-specific expression in synovial tissue of early treatment-naïve RA patients

**DOI:** 10.64898/2026.01.30.26345204

**Authors:** I.C. den Hond, M.J.T. Reinders, M. Lewis, F. Rivellese, C. Pitzalis, R. Knevel, E.B. van den Akker

## Abstract

**Objectives:** Rheumatoid arthritis (RA) exhibits clinical and biological heterogeneity, with synovial tissue stratified into histological pathotypes: lympho-myeloid, diffuse-myeloid, and pauci-immune fibroid. Although GWAS have uncovered RA risk loci, how genetic risk relates to synovial immunopathology remains unclear. To better understand how genetic predisposition may shape divergent early disease mechanisms, we characterised the expression patterns of GWAS-identified RA susceptibility genes and related rheumatic diseases across the synovial pathotypes.

**Methods:** RNA-sequencing data from synovium of 87 treatment-naïve, early RA patients from the Pathobiology of Early Arthritis Cohort. Differential gene expression between pathotypes and pathway enrichment analyses were performed using susceptibility genes for RA, osteoarthritis (OA), ankylosing spondylitis, psoriatic arthritis and systemic lupus erythematosus.

**Results:** RA susceptibility gene expression in synovial tissue separated patients by pathotype and correlated with markers of disease activity. RA susceptibility genes were significantly enriched among genes upregulated in lympho-myeloid synovium and linked to lymphocyte activation and differentiation pathways. In contrast, OA susceptibility genes were upregulated in diffuse-myeloid and fibroid synovium. Both patterns were most pronounced in ACPA-positive and directionally consistent in ACPA-negative patients.

**Conclusion:** RA genetic susceptibility is not evenly distributed across synovial pathotypes but is strongly biased toward the lympho-myeloid pathotype, indicating that current GWAS signals preferentially capture immune-driven disease mechanisms. Enrichment of OA susceptibility genes in diffuse-myeloid and fibroid pathotypes, even among ACPA-positive patients, suggests shared biological features between auto-immune and non-inflammatory degenerative joint diseases in certain RA subtypes. Synovial pathotype stratification is therefore essential for interpreting genetic risk and understanding disease heterogeneity.

**Key messages:** *What is already known on this topic:*

- Rheumatoid arthritis (RA) is clinically and biologically heterogeneous, and its affected synovial tissue can be stratified into distinct immunohistological pathotypes.
- GWAS have identified numerous genetic risk loci for RA and related rheumatic and inflammatory diseases.
- It remains poorly understood how RA genetic risk relates to synovial tissue heterogeneity.

*What this study adds:*

- GWAS-identified RA susceptibility genes show strong, pathotype-specific expression in synovial tissue, with marked enrichment in the lympho-myeloid pathotype.
- OA susceptibility genes are primarily upregulated in diffuse-myeloid and pauci-immune fibroid RA synovium, indicating shared fibroblast- and remodelling-related pathways.
- These gene expression patterns are most pronounced in ACPA-positive RA but remain directionally consistent in ACPA-negative RA.

*How this study might affect research, practice or policy:*

- Synovial pathotype stratification should be incorporated into genetic studies of RA.
- Pathotype-aware genetic studies may improve patient stratification and guide development of more targeted therapeutic strategies.

## Introduction

Both genetic and environmental factors contribute to rheumatoid arthritis (RA) disease aetiology, with genetic predispositions accounting for up to 50% of the overall disease risk^1^. A key clinical factor differentiating RA patients is the presence of anti-citrullinated protein antibodies (ACPA), with ACPA-positive patients having a more aggressive disease course and higher relapse rates. Despite this clear clinical stratification, the molecular mechanisms underlying RA heterogeneity in distinct disease phenotypes remain incompletely understood.

Genome-wide association studies (GWAS) have identified genetic variants predisposing RA, with the latest study reporting 124 loci^2^. The strongest association with RA is found in the human leukocyte antigen (HLA) locus, which explains a substantial proportion of the genetic risk in ACPA-positive patients^3^. In addition, many non-HLA loci are implicated in immune regulatory pathways including T-cell activation and antigen presentation^4–6^. For instance, *PTPN22* (protein tyrosine phosphatase nonreceptor 22), *STAT4* (signal transducer and activator of transcription 4 protein), *CTLA4* (cytotoxic T-lymphocyte antigen 4), and *PADI4* (peptidyl arginine deiminase type IV) have been identified to play a role in RA pathogenesis. However, translation of RA genetic predisposition into a comprehensive understanding of RA aetiology remains challenging. This likely reflects the complexity of genetic regulation, the modest effect sizes of individual single nucleotide polymorphisms (SNPs) on disease risk, and the diverse combinations of variants that RA patients carry^3,7^. Most probably, the heterogeneity in biological mechanisms amongst patients with RA hampers this translation of genetic associations.

Research efforts to disentangle RA heterogeneity have recently expanded to integration of tissue-level molecular characteristics. One promising direction is the histological subtyping of RA synovial tissue into three distinct pathotypes^8,9^: (1) the *lympho-myeloid pathotype*, characterised by a high infiltration of T- and B-lymphocytes; (2) the *diffuse-myeloid pathotype*, dominated by the presence of macrophages but little B-cells; and (3) the *pauci-immune fibroid pathotype*, which shows little to no lymphocyte infiltration and the presence of fibroblasts. Gene expression studies of the biopsied RA synovial tissue show marked differences between pathotypes that can neither be predicted from clinical markers, nor by gene expression in the matching peripheral blood samples^8^. This not only highlights our inability to capture disease heterogeneity in the localized affected tissue using peripheral markers, it also extends the scope of how genetic predispositions may contribute to disease heterogeneity. These distinct synovial pathotypes may provide a new framework to link RA susceptibility genes to specific disease mechanisms more effectively rather than treating RA as a single, homogeneous entity.

Recent work from Galluccio et al. suggested that pauci-immune fibroid synovitis is not unique to RA, as synovial biopsies from patients with other autoimmune or inflammatory diseases, including PsA and juvenile idiopathic arthritis, showed similar histopathological features^10^. The pauci-immune fibroid pathotype has also been observed in synovial tissue of patients with PsA^11^ and OA^12^. These observations indicate that synovial pathotypes may not be strictly confined to current clinical disease classifications and may instead reflect shared or overlapping pathogenic processes across different arthritic conditions. Consequently, certain synovial subsets may share a genetic risk component with other diseases, including systemic lupus erythematosus (SLE), axial spondyloarthritis (Ax-SpA), psoriatic arthritis (PsA), and possibly osteoarthritis (OA). Investigating the expression of GWAS-identified susceptibility genes from these diseases within RA synovial tissue may therefore help to uncover common molecular pathways operating at the tissue level and to refine our understanding of disease heterogeneity beyond conventional diagnostic boundaries.

In this work, we aim to characterise the expression patterns of GWAS-identified RA, OA, SLE, Ax-SpA, and PsA susceptibility genes across synovial pathotypes and explore the relevance of susceptibility genes associated with other rheumatic diseases in RA synovial tissue (Figure 1). By integrating genetic risk signatures with tissue-level synovial pathotypes, our work aims to improve the understanding of molecular heterogeneity within RA synovial tissue and ultimately facilitate improved patient stratification for targeted treatment therapies.

**Figure 1.**
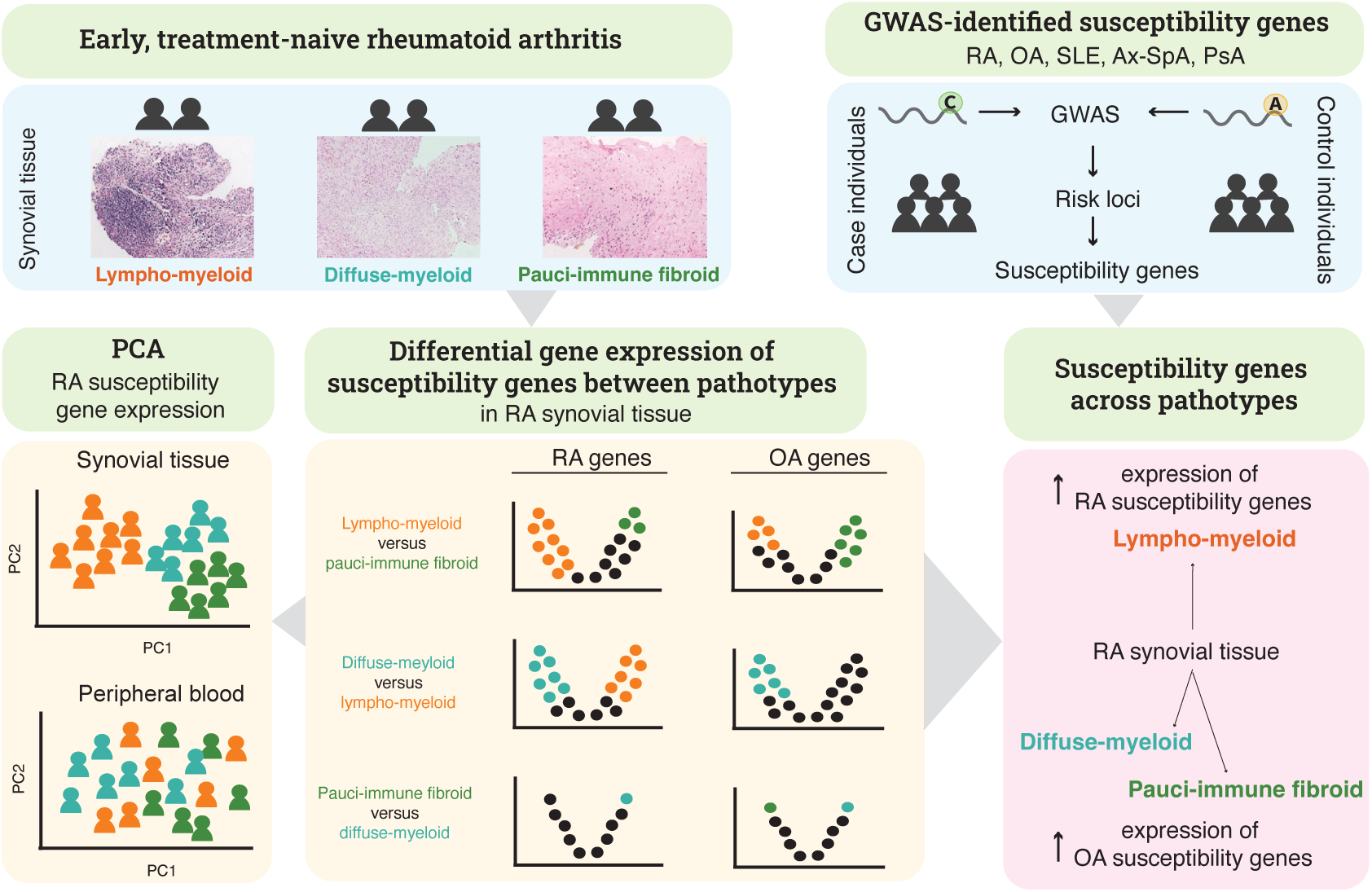
Study overview. Treatment-naïve patients with early rheumatoid arthritis (RA) (symptom duration < 1 year) were stratified into three pathotypes (lympho-myeloid, diffuse-myeloid, pauci-immune fibroid) based on histology and immunohistochemistry profiles of biopsied synovial tissue^8^. The expression of GWAS-identified RA, osteoarthritis (OA), systemic lupus erythematosus (SLE), psoriatic arthritis (PsA), and ankylosing spondylitis (Ax-SpA) susceptibility genes is analysed in synovial tissue and peripheral blood. Principal component analysis on RA susceptibility gene expression in synovial tissue shows that patients tend to group together by pathotype, and differential gene expression in synovial tissue reveals that RA and OA susceptibility genes are not uniformly expressed across pathotypes.

## Methods

### Pathobiology of Early Arthritis Cohort (PEAC)

This study used 87 patients enrolled in the Pathobiology of Early Arthritis Cohort^8,9^ (PEAC) fulfilling 2010 ACR/EULAR RA Classification Criteria. The PEAC cohort^8^ received ethical approval from the UK Health Research Authority (REC 05/Q0703/198, National Research Ethics Service Committee London – Dulwich). All patients gave written informed consent. Based on immunohistochemistry staining of the synovial tissue, Lewis *et al.* have classified each sample (N = 87) as one of three pathotypes: lympho-myeloid (N = 45), diffuse-myeloid (N = 20), or pauci-immune fibroid (N = 16)^8^. Six samples were labelled as ungraded. RNA sequencing was performed on both synovial tissue and peripheral blood. We defined ACPA-positivity as anti-cyclic citrullinated peptide (CCP) equal to or higher than 20 (35 lympho-myeloid patients, 10 diffuse-myeloid patients, 10 pauci-immune fibroid patients). We calculated associations between clinical features and synovial pathotype groups using Fisher’s exact test for categorical variables and one-way analysis of variance (ANOVA) for continuous variables.

### RNA-seq processing

RNA sequencing samples and clinical data were accessed (87 synovium and 67 peripheral blood samples) from ArrayExpress under accession code E-MTAB-6141. 10 single-end sequencing samples from peripheral blood were removed to mitigate batch effects, leaving 57 blood samples. Transcript abundances were estimated over GENCODE v46/GRCh38 transcripts using Kallisto (version 0.48.0) and tximport (version 1.30.0).

### GWAS-identified susceptibility genes

RA susceptibility genes were obtained from Ishigaki *et al*.^2^ (GWAS catalog accession ID GCST90132222), comprising 145 variants (excluding the MHC region) linked to 181 RA susceptibility genes. The GWAS includes 35,871 RA patients and 240,149 control individuals. As this GWAS has excluded the HLA region, we manually added three HLA genes (*HLA-DRB1*, *HLA-B*, and *HLA-DPB1*) previously shown to be associated with ACPA-positive RA^13^. *HLA-DRB1* and *HLA-B* are also associated with ACPA-negative RA^14,15^. OA susceptibility genes were obtained from Boer *et al*.^16^ (GWAS catalog PubMed ID 34450027), where 100 variants are linked to 126 genes. This GWAS includes 649,173 control individuals and 177,517 OA patients. PsA, SLE and Ax-SpA susceptibility genes were obtained from Verma *et al*.^17^. For PsA (GCST90480459) 16 variants have been mapped to 22 genes based on 3,723 cases and 628,576 controls. For SLE (GCST90480453) 10 variants have been mapped to 12 genes, based on 1,965 cases and 630,574 controls. For Ax-SpA (GCST90480502), 25 variants have been mapped to 35 genes. This study included 2,080 patients and 630,112 controls.

### Gene expression processing

Gene expression counts were filtered for minimal expression (at least 10 counts for at least n samples, where n is the smallest group size (16 in synovium and 12 in blood for all patients, 10 in synovium and 4 in blood for ACPA-positive patients, 6 in synovium and 5 in blood for ACPA-negative patients) and normalised using DESeq2’s median of ratios method.

### Principal component analysis

Principal component analysis is performed on the expression of the RA susceptibility genes^2^. Prior to PCA, the gene expression counts are transformed using DESeq2’s (version 1.42.0) variance stabilizing transformation. Gene expression counts from the ungraded samples are used in the principal component analysis (PCA) but not in differential gene expression analysis. Silhouette score (averaged over the classes) is computed to evaluate the separation of pathotype clusters in the two-dimensional PCA space with Euclidean distance as the distance metric.

### Differential gene expression

We performed differential gene expression analysis between pathotypes per tissue type with DESeq2 with sex and pathotype as model covariates for all three pairwise pathotype combinations (lympho-myeloid versus diffuse-myeloid, lympho-myeloid versus pauci-immune fibroid, and diffuse-myeloid versus pauci-immune fibroid samples). Ungraded samples are removed here. Genes are considered differentially expressed with an absolute log fold change > 1 and false discovery adjusted *p* < 0.05.

### Overrepresentation of susceptibility genes in differentially expressed genes

We calculated one-sided Fisher’s exact test to determine whether the set of up- or downregulated RA (or OA) susceptibility genes are overrepresented in the set of total up- or downregulated genes for each comparison of pathotypes per tissue type.

### Overrepresentation of GO biological pathways in the differentially expressed susceptibility genes

We used the enrichGO function from ClusterProfiler (version 4.10.0) to find which gene ontologies (biological processes) are overrepresented among the up- or downregulated RA (or OA) susceptibility genes with as background the set of all up- or downregulated genes, separate for each pairwise comparison of pathotypes.

### Patient and public involvement statement

This work was discussed at multiple SPIDeRR consortium meetings in which several patient research partners participated.

## Results

### Clinical patient characteristics

To investigate the role of GWAS-identified susceptibility genes in synovial rheumatoid arthritis pathotypes, we analysed gene expression profiles derived from ultrasound-guided synovial biopsy samples collected as part of PEAC (Methods)^8^. The patients included in this cohort (N = 87) presented with early rheumatoid arthritis (average disease duration 5.5 +- 3.1 months) and high disease activity (average 28-joint disease activity score 5.9 +- 1.3) (Table 1). The ESR, highest in lympho-myeloid patients, is significantly different between pathotype groups. In addition, there are some trends where the lympho-myeloid patients have a more severe disease activity (DAS28) and a slightly higher frequency of ACPA-positivity than the other patients. In the larger PEAC cohort, ACPA-positivity and DAS28 are significantly higher in the lympho-myeloid patients^9^.

**Table 1.** Baseline clinical features of early RA, treatment-naïve patients from the PEAC cohort for those with synovial biopsy taken. P-values are calculated across pathotypes excluding the ungraded samples with one-way ANOVA for continuous variables and Fisher’s exact for categorical variables (Methods). Values indicate mean +- standard deviation, mean (percentage) or counts (sex). ESR = erythrocyte sedimentation rate, CRP = C-reactive protein, CCP = cyclic citrullinated protein antibody, RF = rheumatoid factor, VAS = visual analogue scale, TJC = tender joint count, SJC = swollen joint count, HAQ = health assessment questionnaire score, DAS28 = 28-joint disease activity score.

| *All patients* | *All* | *Lympho-myeloid* | *Diffuse-myeloid* | *Pauci-immune fibroid* | *p-value* | *Ungraded* |
| --- | --- | --- | --- | --- | --- | --- |
| *n* | 87 | 45 | 20 | 16 |  | 6 |
| *age (years)* | 54 +- 17 | 54 +- 16 | 54 +- 20 | 56 +- 16 | 0.9 | 47 +- 12 |
| *sex (female/male)* | 64 / 23 | 34 / 11 | 14 / 6 | 12 / 4 | 0.94 | 4 / 2 |
| *disease duration (months)* | 5.9 +- 3.2 | 5.7 +- 3.3 | 5.0 +- 3.0 | 6.5 +- 3.3 | 0.4 | 7.8 +- 2.5 |
| *ESR (mm/hr)* | 45 +- 29 | 54 +- 30 | 36 +- 25 | 35 +- 31 | 0.022 | 33 +- 18 |
| *CRP (ug/mL)* | 23 +- 33 | 27 +- 27 | 17 +- 25 | 25 +- 56 | 0.59 | 7.3 +- 7 |
| *ACPA positive (%)* | 56 (64%) | 35 (78%) | 10 (50%) | 10 (63%) | 0.072 | 1 (17%) |
| *CCP (U/mL)* | 220 +- 218 | 262 +- 208 | 134 +- 177 | 255 +- 251 | 0.07 | 105 +- 243 |
| *RF positive (%)* | 62 (71%) | 36 (80%) | 14 (70%) | 10 (62.5%) | 0.33 | 2 (33.33%) |
| *VAS* | 65 +- 23 | 67 +- 24 | 62 +- 23 | 59 +- 26 | 0.53 | 78 +- 16 |
| *TJC* | 12 +- 8 | 13 +- 7 | 10 +- 6 | 12 +- 9 | 0.27 | 17 +- 8 |
| *SJC* | 8 +- 6 | 9 +- 6 | 7 +- 4 | 7 +- 7 | 0.27 | 9 +- 6 |
| *HAQ* | 1.55 +- 0.76 | 1.63 +- 0.80 | 1.37 +- 0.55 | 1.59 +- 0.82 | 0.44 | 1.5 +- 0.96 |
| *DAS28* | 5.9 +- 1.3 | 6.2 +- 1.3 | 5.5 +- 1.1 | 5.4 +- 1.6 | 0.078 | 6.4 +- 1.3 |

### Patterns of RA susceptibility gene expression vary across pathotypes

In the most recent and largest RA GWAS, Ishigaki *et al*. described 181 RA susceptibility genes^2^. Including *HLA-DRB1*, *HLA-B*, and *HLA-DPB1* for their known association with RA^13–15^, the final RA susceptibility gene set consists of 184 genes. Of these, 143 were expressed in synovial tissue, and 137 in peripheral blood (Figure 2A). Principal component analysis (PCA) of expressed RA susceptibility genes in synovial tissue reveals that patients tend to cluster by histologically defined pathotypes (Figure 2B). Principal components (PC) 1 and 2 capture the transcriptomic difference between lympho-myeloid, diffuse-myeloid, and pauci-immune fibroid synovial tissue. PC1 is negatively correlated with clinical features that indicate disease severity such as ESR, DAS28, CRP, and CCP, indicating a more inflammatory disease state in the lympho-myeloid patient subgroup. PC2 on its own highlights differences between pauci-immune fibroid and diffuse-myeloid synovial tissue, with CRP being higher in pauci-immune fibroid patients (Figure 2C). This suggests that the gene expression patterns of RA susceptibility genes reflect underlying molecular differences between these pathotypes. In contrast to synovial tissue, no clear separation of samples by pathotype was observed in peripheral blood based on expression of RA susceptibility genes (Supplementary Figure S1, Supplementary Table S1). Together, these observations indicate that the expression signature of the RA susceptibility genes in synovial tissue captures the transcriptomic differences between synovial pathotypes.

**Figure 2.**
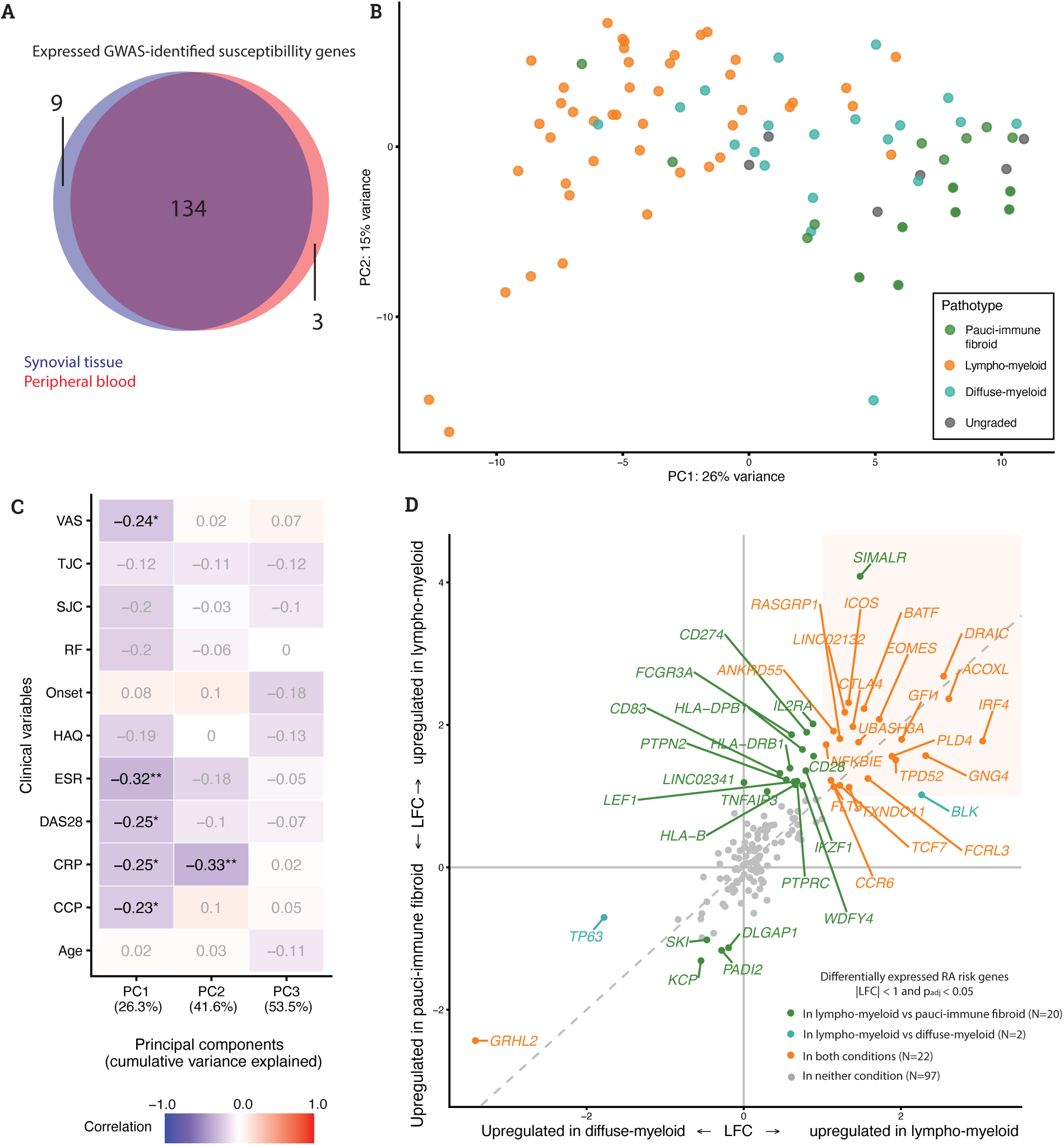
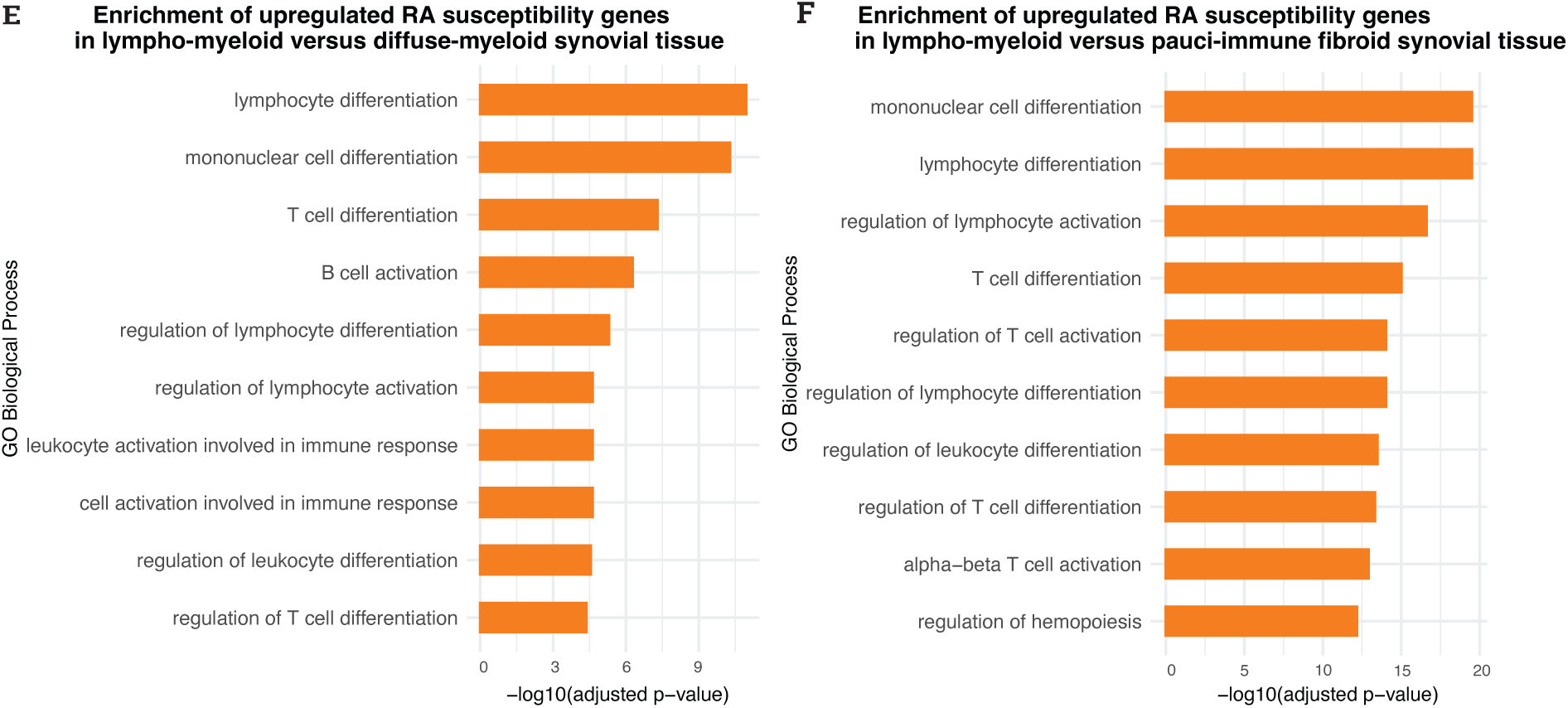
Expression of RA susceptibility genes in synovial tissue of early RA patients. **A)** Overlap between expressed RA susceptibility genes in synovial tissue (N = 143) and peripheral blood (N = 137). **B**) Principal component analysis of RA susceptibility gene expression (N = 143) in synovial tissue showing a distinction between lympho-myeloid, diffuse-myeloid and pauci-immune fibroid patients on principal component (PC) 1 and 2. **C)** Spearman’s correlation between clinical variables and PC1, 2 and 3. PC1, 2 and 3 are calculated on expression of RA susceptibility genes in synovial tissue (as shown in **B**). **D**) Effect size (log fold change) of differentially expressed RA susceptibility genes in diffuse-myeloid versus lympho-myeloid synovial tissue (x-axis) and pauci-immune fibroid versus lympho-myeloid synovial tissue (y-axis) in synovial tissue of early RA patients. Genes in the orange square are upregulated in lympho-myeloid synovial tissue compared to both diffuse-myeloid and pauci-immune fibroid synovial tissue. The diagonal line indicates y = x. LFC = log fold change. **E, F**) Gene ontology (GO) biological processes that are overrepresented among the RA susceptibility genes that are upregulated in lympho-myeloid versus diffuse-myeloid (**E**) and pauci-immune fibroid (**F**) synovial tissue.

### RA susceptibility genes are upregulated in lympho-myeloid synovial tissue

To pinpoint the transcriptomic differences of RA susceptibility genes between pathotypes, we conducted differential gene expression (DGE) analyses (Methods). DGE on all 24,404 genes expressed in synovial tissue revealed substantial numbers of differentially expressed genes for the lymphoid- versus diffuse-myeloid, and the lymphoid- versus pauci-immune fibroid pathotype contrasts. This indicates broad transcriptional differences between the pathotypes (Figure 3A, 3B), in line with previous results from Lewis *et al*.^8^.

**Figure 3.**
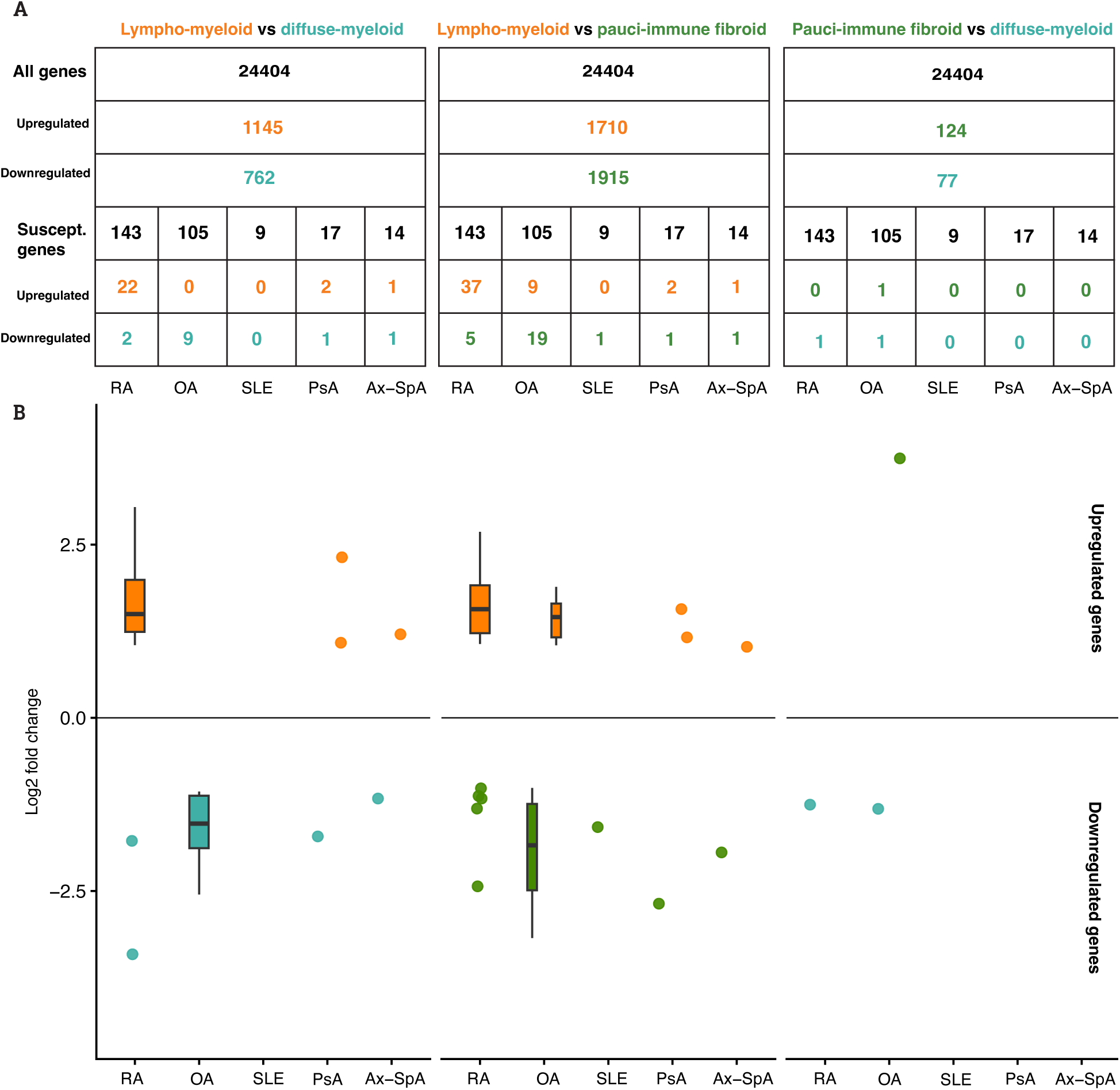
Differentially expressed genes between pathotypes in synovial tissue of early, treatment-naive RA patients.**A)** Number of expressed and differentially expressed genes between each pair of pathotypes in synovial tissue of RA patients, including the number of expressed and differentially expressed genes that belong to the RA, OA, SLE, PsA, and Ax-Spa susceptibility gene set. Asterisk indicates significant (p < 0.05) overrepresentation (Fisher’s exact test, Methods) of differentially expressed susceptibility genes given the total number of upregulated genes for that disease. **B)** Log fold changes of differentially expressed susceptibility genes for all diseases and all pathotype comparisons. Boxes are plotted if more than 5 genes are differentially expressed, and show mean and standard deviation. A blank column indicates that no genes are differentially expressed for that disease and that pathotype comparison.

We observed that a shared subset of the RA susceptibility genes is upregulated in lympho-myeloid synovial tissue, in contrast to both diffuse-myeloid (N = 22) and pauci-immune fibroid synovial tissue (N = 37) (Figure 3A, 3B), including well-known RA susceptibility genes such as *CTLA4, CCR6, and IRF4*. These upregulated genes were more often upregulated than expected (Fisher’s exact test, Methods) relative to the number of upregulated genes in lympho-myeloid synovial tissue, both in the diffuse-myeloid (*p* = 9e-9) and pauci-immune fibroid (*p* = 2e-10) contrast. This upregulation shows a similar effect size (log fold change) between lympho-myeloid synovial tissue and each of the other two pathotypes (Figure 2D). Two other genes are upregulated in the diffuse-myeloid (*TP63*), or in both diffuse-myeloid and pauci-immune fibroid (*GRHL2*) synovial tissue contrasted to lympho-myeloid synovial tissue. This shared gene expression signature distinguishes the lympho-myeloid pathotype from both the fibroid- and diffuse-myeloid pathotype and suggests that the genetic risk for RA is most prominently reflected in the lympho-myeloid pathotype.

### OA susceptibility genes are upregulated in diffuse-myeloid and pauci-immune fibroid RA synovial tissue

Given that the RA susceptibility genes seem to capture only part of the synovial heterogeneity, we were interested whether GWAS-identified susceptibility genes of other rheumatic diseases like OA, PsA, SLE, and Ax-SpA could explain expression differences between pathotypes. The OA susceptibility gene set (104 genes expressed in synovial tissue) shares two genes (*HLA-DPB1* and *LINC02341*) with the RA susceptibility gene set (Supplementary Figure S4). The effect of upregulation of OA susceptibility genes is opposite of how the RA susceptibility genes behave across pathotypes: a subset of the OA susceptibility genes is upregulated in the synovial tissue of both diffuse-myeloid and pauci-immune fibroid RA patients, compared to the synovial tissue of lympho-myeloid RA patients (Figure 3A, 3B). The effect in both comparisons is driven by the same set of OA susceptibility genes, reflected in similar effect sizes for these genes in the comparison between diffuse-myeloid and pauci-immune fibroid synovial tissue on one hand, and lympho-myeloid synovial tissue on the other hand (Figure 4A). Overrepresentation analysis showed that OA susceptibility genes are more often upregulated than expected in diffuse-myeloid (*p* = 0.000804) and pauci-immune fibroid (*p* = 0.00153) synovial tissue in contrast with lympho-myeloid synovial tissue.

**Figure 4.**
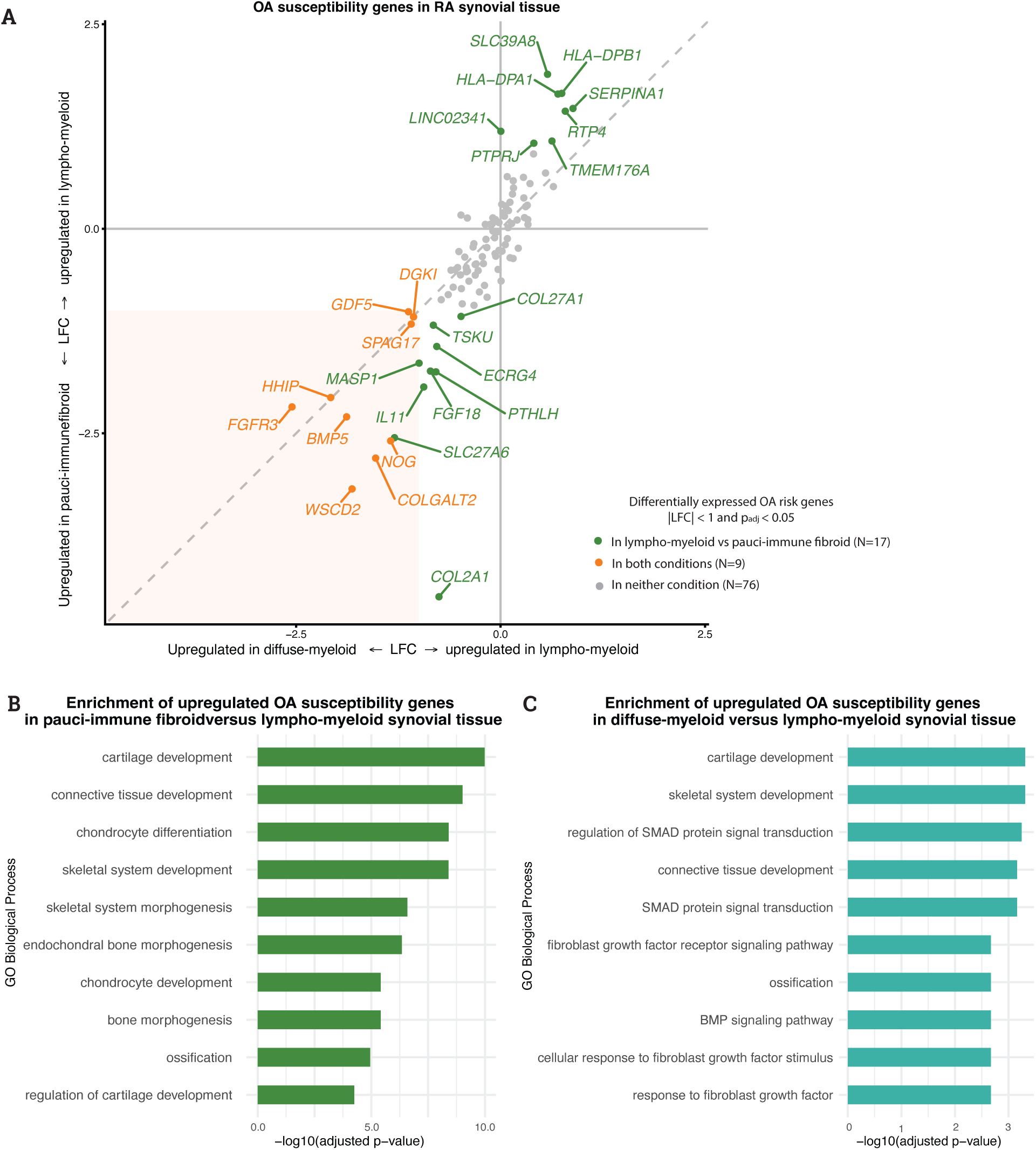
Expression of OA susceptibility genes in RA synovial tissue. **A)** Effect size (log fold change) of differentially expressed OA susceptibility genes in diffuse-myeloid versus lympho-myeloid synovial tissue (x-axis) and pauci-immune fibroid versus lympho-myeloid synovial tissue (y-axis) in synovial tissue of early RA patients. Genes in the orange square are upregulated in both diffuse-myeloid and pauci-immune fibroid synovial tissue compared to lympho-myeloid synovial tissue. The diagonal line indicates y = x. LFC = log fold change. **B, C)** Gene ontology (GO) biological processes that are overrepresented among the OA susceptibility genes that are upregulated in pauci-immune fibroid (**B**) and diffuse-myeloid (**C**) versus lympho-myeloid synovial tissue.

Similar to OA, we analysed the expression of GWAS-identified susceptibility genes for SLE (N = 35), PsA (N = 22), and Ax-SpA (N = 12) in peripheral blood and synovial tissue. These GWAS were based on smaller patient cohorts than those for RA and OA and reported only a small number of risk variants with little overlap between diseases (Supplementary Figure S4). We found that these susceptibility genes were expressed in synovial tissue of RA patients, but not more often than expected differentially expressed between pathotypes (Figure 3A, 3B).

### Enhanced transcriptomic differences between pathotypes amongst ACPA-positive patients

To exclude the possibility of misclassification, we repeated the analysis in the ACPA-positive RA patients of this cohort (N = 56, clinical characteristics in Supplementary Table S2). The signal of upregulation of RA susceptibility genes was more pronounced than seen before in the full patient cohort (Figure 5, Supplementary Table S3). When evaluating pathotype separability only in ACPA-positive patients using a two-dimensional PCA space, the pathotype separation (silhouette score = 0.17, Supplementary Figure S2) was more distinct than in the full patient cohort (silhouette score = 0.11). In the smaller group of ACPA-negative patients, most of the genes did not reach statistical significance for differential expression (Supplementary Table S4). Effect size estimates (log fold changes) were generally smaller for this contrast, yet effect directions were generally consistent between ACPA-positive and ACPA-negative patients. Similarly for the OA susceptibility genes, we found that the effect of upregulation was strongest in the ACPA-positive patient cohort (Supplementary Table S5, Supplementary Figure S3), again showing that OA susceptibility genes are mainly upregulated in diffuse-myeloid and pauci-immune fibroid compared to lympho-myeloid RA synovial tissue.

**Figure 5.**
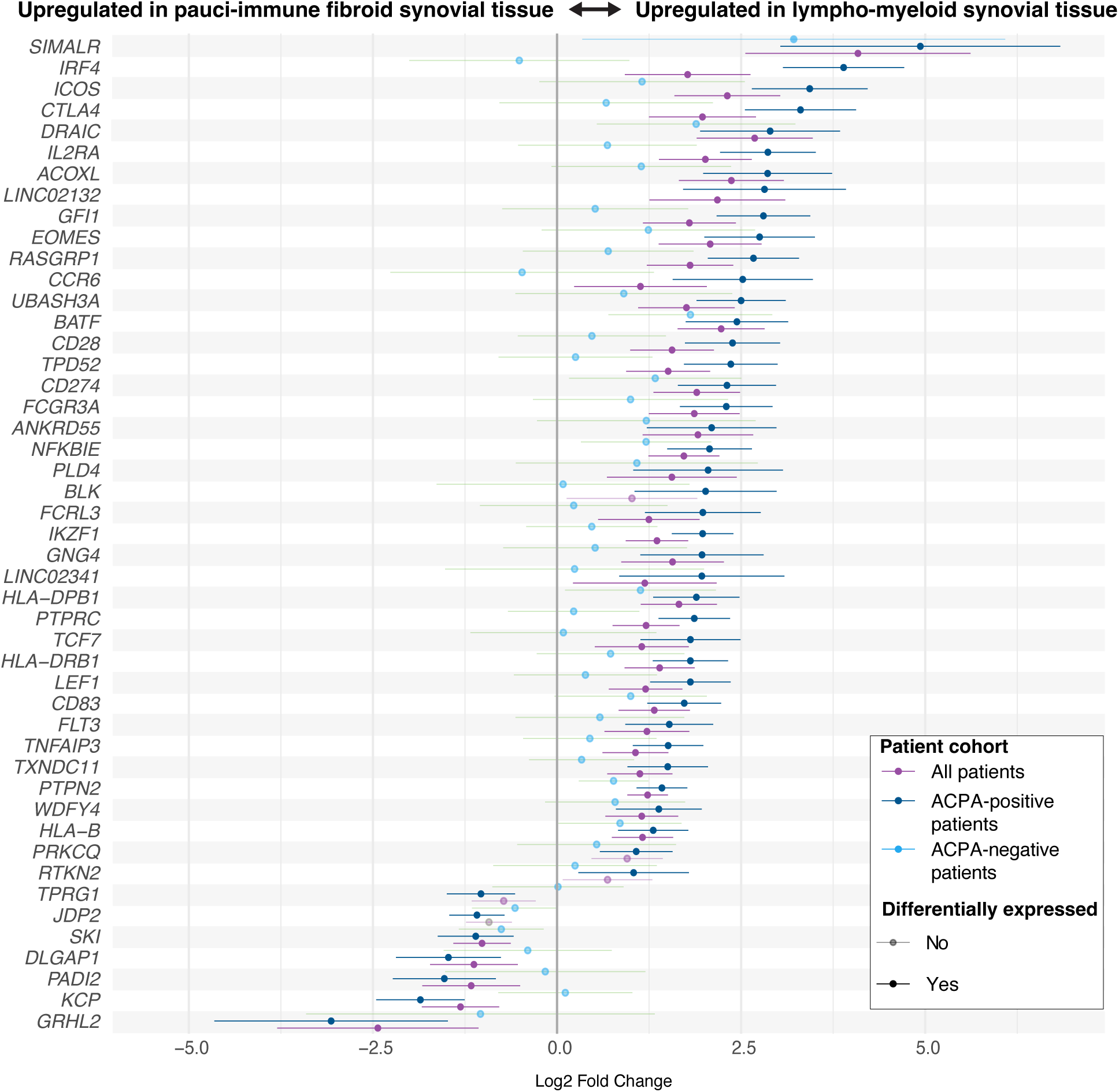
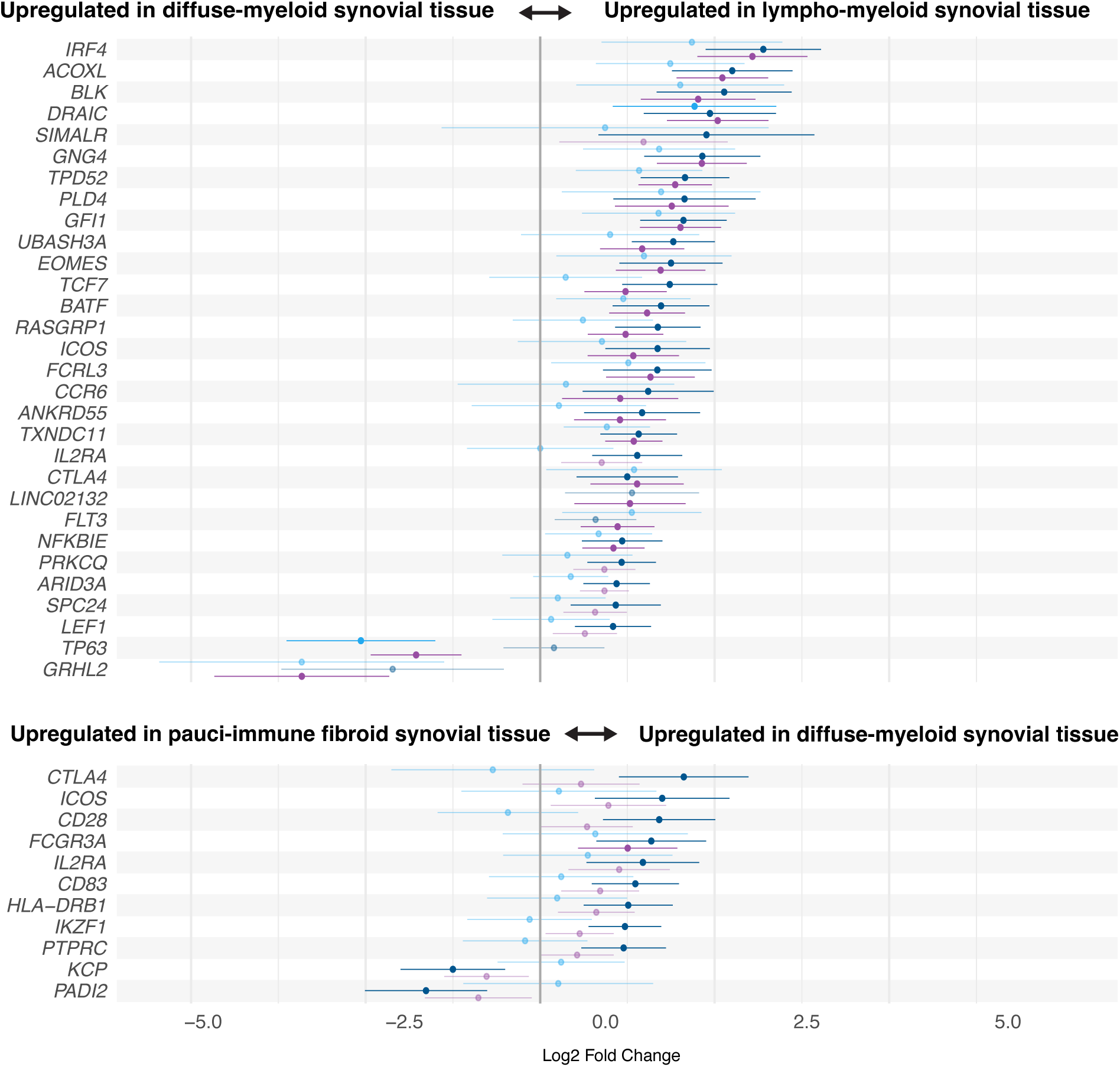
Effect sizes (log fold changes) of differentially expressed RA susceptibility genes in different patient cohorts (ACPA-positive only, ACPA-negative only, all patients) between **A)** lympho-myeloid and diffuse-myeloid, **B)** lympho-myeloid and pauci-immune fibroid, and **C)** pauci-immune fibroid and diffuse-myeloid synovial tissue. A gene is shown if it is significantly differentially expressed in at least one patient cohort. Horizontal lines indicate 95% confidence intervals for log fold change.

### Transcriptomic differences of susceptibility genes reflect pathotype-specific immune and tissue-remodelling pathways

Next, we aimed to better understand how transcriptomic differences between pathotypes might impact tissue function using Gene Ontology (GO) gene set enrichment analyses. First, we performed gene set enrichment analysis on the RA susceptibility genes that were upregulated in lympho-myeloid in comparison to diffuse-myeloid or pauci-immune fibroid synovial tissue. Many of the upregulated RA susceptibility genes in lympho-myeloid synovial tissue are involved in regulating lymphocyte development and activation (Figure 2E and 2F), which is in line with the lympho-myeloid pathotype showing high lymphocyte infiltration. For instance, upregulation of genes like *CTLA4*, *ICOS*, *IRF4*, and *CCR6* is consistent with the high level of immune cell infiltration in lympho-myeloid synovial tissue, as these genes are key regulators of T-cell activation and differentiation. Consistent with the DGE analyses, gene set enrichment analyses were highly similar for the lymphoid- versus diffuse-myeloid, and the lymphoid- versus pauci-immune fibroid pathotype contrasts. The only notable difference was the ‘B-cell activation’ gene set for the lymphoid- versus diffuse-myeloid contrast, most probably related to the differentially expressed *BLK* gene in only the diffuse-myeloid condition. Second, we performed gene set enrichment analysis on the OA susceptibility genes that were upregulated in diffuse-myeloid or pauci-immune fibroid synovial tissue. The enrichment of connective tissue remodelling and growth factor receptor signalling pathways within the OA susceptibility gene set (Figure 4B and 4C) indicates active processes of tissue repair and pathological remodelling. This aligns with the cellular composition of especially the RA pauci-immune fibroid synovial pathotype, which is dominated by fibroblasts. The upregulation of OA susceptibility genes in specific subsets of RA synovial tissue suggests shared pathogenic mechanisms between OA and pauci-immune fibroid/diffuse-myeloid RA, particularly in bone and extracellular matrix remodelling. Immune activation dominates the lympho-myeloid pathotype, whereas fibroblast-driven tissue remodelling pathways are more prominent in diffuse-myeloid and pauci-immune fibroid pathotypes, with potential overlap between RA and OA mechanisms.

## Discussion

This study presents a comprehensive analysis linking GWAS-identified susceptibility genes of RA and related rheumatic diseases to molecular pathotypes previously identified in RA synovial tissue, thereby connecting genetic risk to tissue-level heterogeneity. RA is increasingly recognised as a clinical and biological heterogeneous disease, yet most genetic studies have treated RA as a single entity. Our results demonstrate that this heterogeneity is reflected at the transcriptomic level with RA susceptibility genes being non-uniformly expressed across synovial pathotypes, with the current knowledge on RA genetic architecture being biased towards the lympho-myeloid pathotype.

Specifically, RA susceptibility genes were predominantly upregulated in the lympho-myeloid synovial pathotype, supporting the notion that current RA genetic risk mainly maps to lymphoid-driven disease mechanisms. In line with the described biology of the lympho-myeloid pathotype, the upregulated genes are primarily involved in adaptive immune responses at the site of inflammation, most notably T-cell activation and regulation. In particular, the differential expression of cytotoxic T lymphocyte-associated protein 4 (*CTLA4*) seems relevant, as this gene is expressed in T regulatory cells and suppresses T cell activation via inhibitory signalling^18^. Its central role in RA is therapeutically targeted by Abatacept, which mimics CTLA-4 by inhibiting T-cell activation^19^. In contrast, relatively few RA susceptibility genes were upregulated in diffuse-myeloid or pauci-immune fibroid synovial tissue. One notable exception is *TP63*, a putative methotrexate target^20^, which was selectively upregulated in diffuse-myeloid synovium relative to the lympho-myeloid pathotype.

Strikingly, several of the RA susceptibility genes showing the most pronounced up- or downregulation in lympho-myeloid RA synovial tissue, are long non-coding RNAs that have not previously been linked to established RA disease mechanisms. In our analysis, *SIMALR, DRAIC, and LINC02132* were found to be strongly upregulated in lympho-myeloid RA synovial tissue, suggesting pathotype-specific regulatory roles. *SIMALR* has recently been reported as a marker of M1 inflammatory macrophages^21^, implying that its elevated expression may contribute to the sustained immune cell activation and inflammatory signalling that define the lympho-myeloid synovial pathotype. Similarly, both *DRAIC* and *LINC02132 h*ave been reported in the regulation of immune responses in the contexts of cancer^22^, and autoimmune diseases^23^, but have not previously been associated with RA pathogenesis. These results suggest that these lncRNAs may represent previously unrecognized molecular regulators within RA synovium, particularly in the immune-dominant lympho-myeloid pathotype. In sharp contrast, the transcription factor *GRHL2* was consistently upregulated in both diffuse-myeloid and pauci-immune fibroid RA synovial tissue, highlighting a distinct stromal-associated genetic signal. Recently, Ge *et al*. have identified a potential functional relationship between the *GRHL2* RA risk variant and the regulatory landscape of synovial fibroblasts^24^, underscoring the contribution of stromal cells to the pauci-immune fibroid synovial tissue. As *GRHL2* expression is enriched specifically in the pathotypes characterised by stromal expansion and tissue remodelling, this further supports the notion that RA genetic risk variants could preferentially act in specific RA pathotypes. Together, this suggests that the RA susceptibility genes showing strongest up- or downregulation across synovial pathotypes are not just the classical immune mediators, but include lncRNAs whose function remains largely unexplored in RA.

As the lympho-myeloid pathotype can be described with the upregulation of the RA susceptibility genes, we investigated whether the genetic predisposition of the diffuse-myeloid and pauci-immune fibroid pathotype could be better understood by examining the effects of GWAS-identified susceptibility genes from related rheumatic diseases. Notably, RA synovial tissue from the diffuse-myeloid and pauci-immune fibroid pathotypes instead showed enriched upregulation of OA susceptibility genes. OA is increasingly seen as a disease of imbalanced tissue repair and extracellular matrix remodelling rather than cartilage degeneration. These biological processes also characterise the pauci-immune fibroid RA pathotype. The enrichment of OA risk genes in RA pauci-immune fibroid synovial tissue perhaps reflects convergent fibroblast-driven tissue remodelling programs. These patients may benefit from other types of treatments, as a recent study by Galluccio *et al*. reported that patients with pauci-immune fibroid synovium achieve clinical remission after treatment with a JAK inhibitor^10^. These insights contribute to our understanding of RA heterogeneity and in the future may guide more precise patient stratification and targeted therapeutic approaches.

The persistence of the effect of upregulation of OA susceptibility genes in the ACPA-positive subset indicates that it cannot be explained by serostatus alone. This pattern may reflect an inherent bias in the RA GWAS itself, where most included patients will belong to the lympho-myeloid RA pathotype, and as a result genetic associations are likely dominated by mechanisms active in the lympho-myeloid form of RA. Consequently, these GWAS may be less sensitive to genetic influences affecting the pauci-immune fibroid and diffuse-myeloid pathotypes. In contrast, OA susceptibility genes may reflect pathogenic processes more characteristic to fibroid- and diffuse-myeloid synovial tissue, including extracellular matrix remodelling and fibroblast activation. By comparison, GWAS-identified susceptibility genes for the related immune-mediated rheumatic diseases PsA, SLE, and Ax-SpA were expressed but not differentially regulated across RA pathotypes, likely owing to the smaller number of risk loci reported for these diseases. Recent work from Goldmann *et al*. on the PEAC cohort has highlighted the value of investigating genetic regulation directly in diseased synovial tissue^25^. Their study identified several significant tissue-specific expression quantitative trait loci (eQTL) in synovial tissue linked to RA, OA, and other autoimmune diseases. This provides compelling evidence that the functional consequences of genetic risk loci should be studied within the relevant, affected tissue. However, curiously, the RA susceptibility genes that Goldmann *et al.* identified through synovial eQTL mapping largely differ from those derived from the most recent RA GWAS. TPRA1 is the only gene reported by both Goldmann *et al.* and Ishigaki *et al.*, and notably, it is not differentially expressed between synovial pathotypes in our analysis. This discrepancy can be explained by several aspects, foremost, a lack of statistical power due to a limited availability of available synovial biopsies. Also, differences in methodology are likely to contribute to this discrepancy. For instance, variant-gene assignments remain challenging and underscore the need for the integration of tissue-specific regulatory effects with large-scale genetic association studies.

Differences in gene expression between pathotypes were most prominent when limiting our analyses to ACPA-positive patients. This finding has several implications. First, it emphasizes that differences between pathotypes are identified irrespective of ACPA status, and thus that differences in gene expression cannot be solely attributed to a differential distribution of ACPA-positive patients across pathotypes. Secondly, since ACPA-positivity is highly specific to rheumatoid arthritis patients, our finding also underscores that differences between pathotypes cannot be attributed to misclassification of patients with rheumatic complaints. Lastly, analyses in the smaller ACPA-negative subgroup showed directionally consistent, yet overall weaker effect sizes, as compared to the ACPA-positive subgroup. This holds true for both the upregulation of RA susceptibility genes in lympho-myeloid synovial tissue, as for the upregulation of OA susceptibility genes in pauci-immune fibroid and diffuse-myeloid synovial tissue. We can only speculate why ACPA-negative patients have less pronounced molecular differences between pathotypes, as compared to ACPA-positive patients, but it might point to a larger heterogeneity amongst ACPA-negative patients. Nevertheless, collectively this indicates that the transcriptional patterns of RA and OA susceptibility genes are consistent across relevant patient strata and cannot be attributed to patient misclassification.

This work provides evidence that genetic risk architecture is not equally distributed across synovial pathotypes of early, treatment-naive RA patients, with GWAS-identified RA susceptibility genes being preferentially upregulated in the lympho-myeloid synovial pathotype. These findings highlight the need for pathotype-aware studies to better understand RA disease heterogeneity and genetic risk and warrant future research into the biological mechanisms contributing to a divergent immunopathology of rheumatic disease.

## Data Availability

All data used are available online at ArrayExpress under accession code E-MTAB-6141.

https://www.ebi.ac.uk/biostudies/ArrayExpress/studies/E-MTAB-6141?query=E-MTAB-6141

## Acknowledgements

This project has received funding from Horizon Europe programme under grant agreement no. 101080711 (SPIDeRR), ZonMW klinische fellow no. 40-00703-97-19069, and ZonMw Open Competitie 2021 no. 09120012110075. We acknowledge the PEAC investigators, and PEAC was supported by funding from the UK Medical Research Council (MRC) (grant number G0800648), by funding from the National Institute for Health Research (NIHR) for infrastructural support to the Barts Biomedical Research Centre (BRC) (grant number: NIHR203330), and by Arthritis UK funding for the Experimental Arthritis Treatment Centre (grant number: 20022). Part of the results of this paper were presented at EULAR 2025 under abstract number OPO155 (https://doi.org/10.1016/j.ard.2025.05.176).

## Notes

### Competing Interest Statement

The authors have declared no competing interest.

### Author Declarations

Source data were openly available to the public before initiation of the study. RNA sequencing samples and clinical data were accessed (87 synovium and 67 peripheral blood samples) from ArrayExpress under accession code E-MTAB-6141 GWAS studies can be obtained from GWAS catalog accession IDs: GCST90132222, Pubmed ID 34450027, GCST90480459, GCST90480453, GCST90480502 The PEAC cohort received ethical approval from the UK Health Research Authority (REC 05/Q0703/198, National Research Ethics Service Committee London - Dulwich). All patients gave written informed consent.

