## Supplementary Data for "Rheumatoid arthritis susceptibility genes show pathotype-specific expression in synovial tissue of early treatment-naïve RA patients"

**Supplementary information**

**Supplementary Figure S1 – RA susceptibility genes in peripheral blood of RA patients**

Principal component analysis of expression of RA susceptibility genes in RA peripheral blood

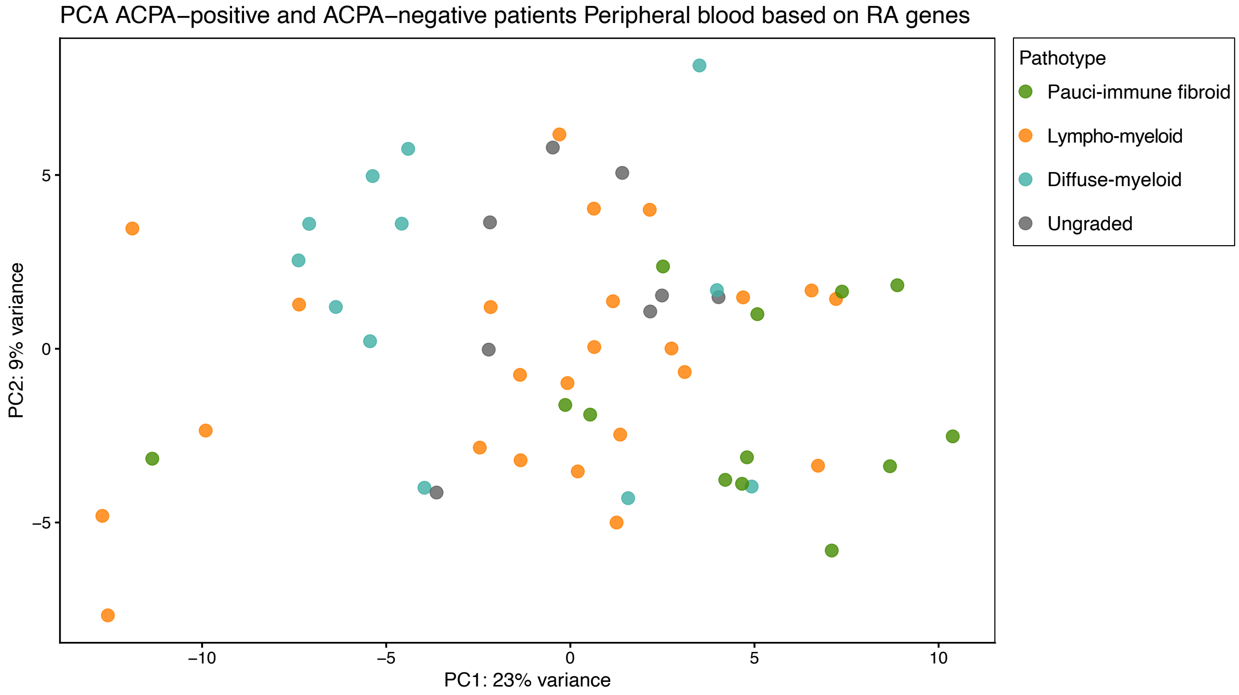


**Supplementary Figure S2 – RA susceptibility genes in RA synovial tissue (ACPA-positive patients)**

Principal component analysis of expression of RA susceptibility genes in RA synovial tissue

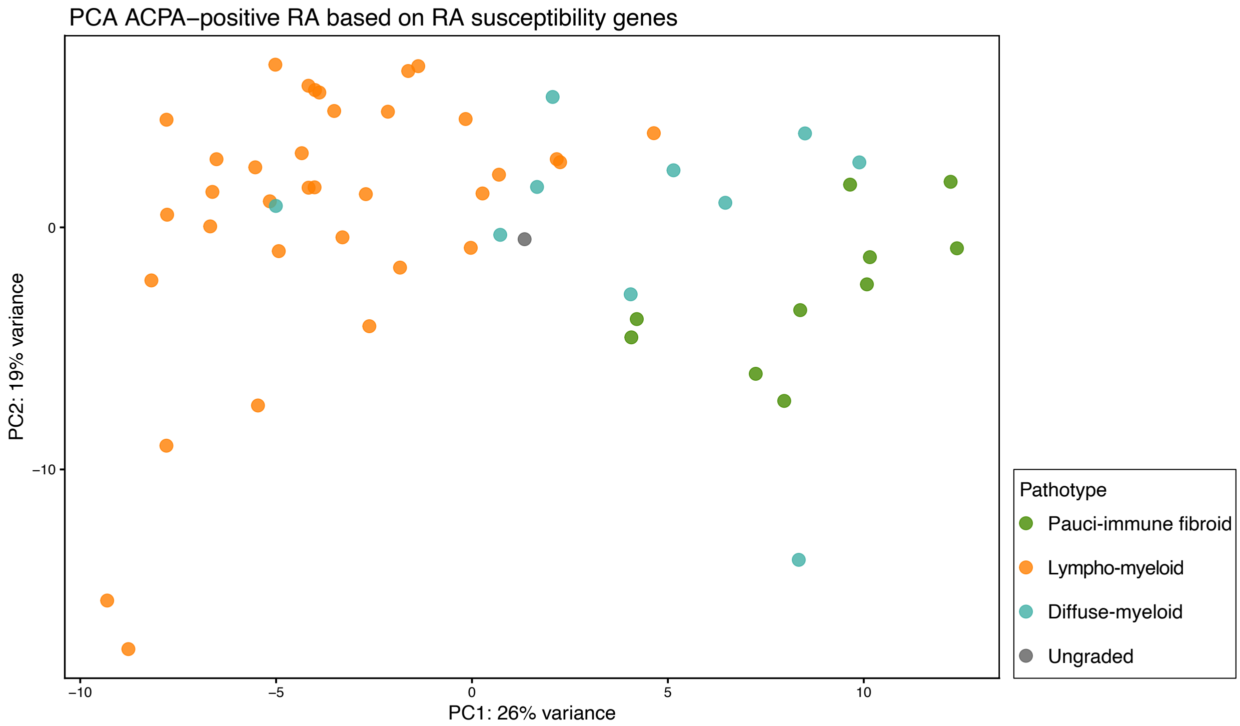


**Supplementary Figure S3 - Effect sizes (log fold changes) of differentially expressed OA susceptibility genes in different patient cohorts (ACPA-positive only, ACPA-negative only, all patients) between A) lympho-myeloid and diffuse-myeloid, B) lympho-myeloid and pauci-immune fibroid, and C) pauci-immune fibroid and diffuse-myeloid synovial tissue. A gene is shown if it is significantly differentially expressed in at least one patient cohort. Horizontal lines indicate 95% confidence intervals for log fold change.**

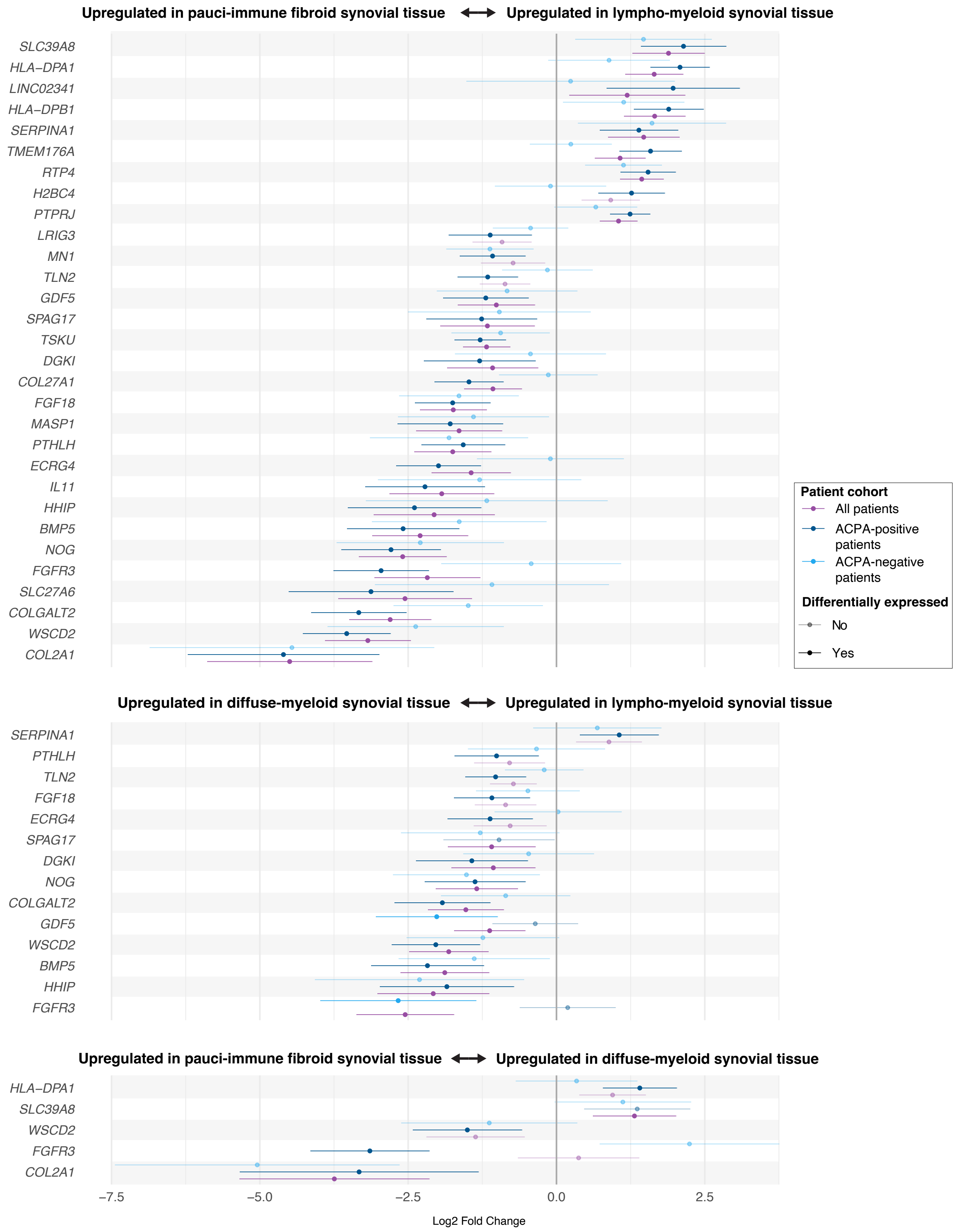
Upregulated in pauci-immune fibroid synovial tissue
Upregulated in lympho-myeloid synovial tissue
SLC39A8
HLA–DPA1
LINC02341
HLA–DPB1
SERPINA1
TMEM176A
RTP4
H2BC4
PTPRJ
LRIG3
MN1
TLN2
GDF5
SPAG17
TSKU
DGKI
COL27A1
FGF18
MASP1
PTHLH
ECRG4
IL11
HHIP
BMP5
NOG
FGFR3
SLC27A6
COLGALT2
WSCD2
COL2A1
Patient cohort
All patients
ACPA-positive patients
ACPA-negative patients
Differentially expressed
No
Yes
Upregulated in diffuse-myeloid synovial tissue
Upregulated in lympho-myeloid synovial tissue
SERPINA1
PTHLH
TLN2
FGF18
ECRG4
SPAG17
DGKI
NOG
COLGALT2
GDF5
WSCD2
BMP5
HHIP
FGFR3
Upregulated in pauci-immune fibroid synovial tissue
Upregulated in diffuse-myeloid synovial tissue
HLA–DPA1
SLC39A8
WSCD2
FGFR3
COL2A1
−7.5
−5.0
−2.5
0.0
2.5
Log2 Fold Change

25

**Supplementary figure S4 – Number of GWAS-identified susceptibility genes that overlap between RA, OA, SLE, PsA, and Ax-SpA. No number indicates no overlap between the susceptibility gene sets of those diseases.**

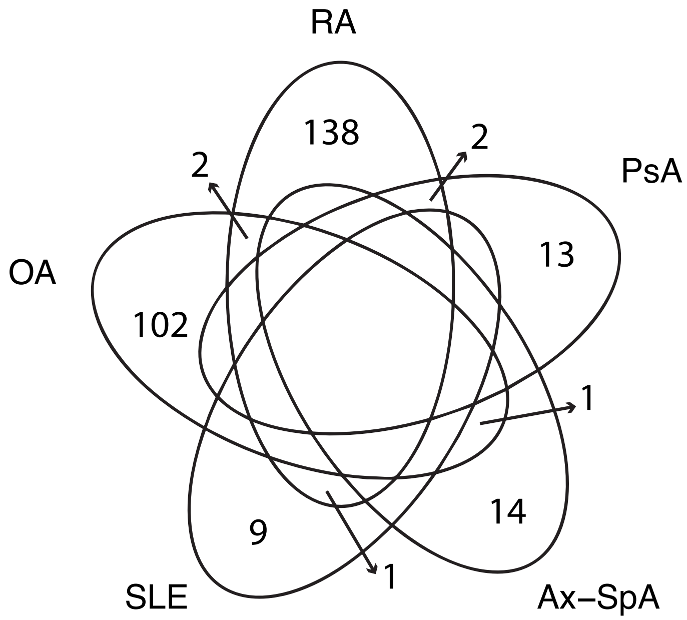

**Supplementary Table S1** – Number of differentially expressed genes between pathotypes in peripheral blood of RA patients, including number of upregulated genes that belong to the RA or OA risk gene set. p-value represents overrepresentation (Fisher's exact test, Methods) of upregulated RA or OA risk genes given the total number of upregulated genes. n.s. = not significant ($p > 0.05$), L = lympho-myeloid, M = diffuse-myeloid, F = pauci-immune fibroid.

| ***Blood*** | **All genes (N = 18777)** | **RA risk genes (N = 137)** | **OA risk genes (N = 74)** |
|---|---|---|---|
| *Upregulated genes in* | | | |
| *L vs M* | 20 | 0<br>n.s. | 0<br>n.s. |
| *M vs L* | 0 | 0<br>n.s. | 0<br>n.s. |
| *L vs F* | 0 | 0<br>n.s. | 0<br>n.s. |
| *F vs L* | 0 | 0<br>n.s. | 0<br>n.s. |
| *F vs M* | 25 | 2<br>p = 0.0142 | 0<br>n.s. |
| *M vs F* | 6 | 0<br>n.s. | 0<br>n.s. |

**Supplementary Table S2** – Clinical features of early RA, treatment-naïve ACPA-positive patients from the PEAC cohort for those with synovial biopsy taken. P-values are calculated across pathotypes excluding ungraded samples with Anova for continuous variables and Fisher's exact for categorical variables (sex, CCP positive, RF positive). Values indicate mean +- standard deviation. ESR = erythrocyte sedimentation rate, CRP = C-reactive protein, CCP = cyclic citrullinated protein antibody, RF = rheumatoid factor, VAS = visual analogue scale, TJC = tender joint count, SJC = swollen joint count, HAQ = health assessment questionnaire score, DAS28 = 28-joint disease activity score.

| ***ACPA-positive patients*** | ***All*** | ***Lympho-myeloid*** | ***Diffuse-myeloid*** | ***Pauci-immune fibroid*** | ***p-value*** | ***Ungraded*** |
|---|---|---|---|---|---|---|
| ***n*** | 56 | 35 | 10 | 10 | | 1 |
| ***age (years)*** | 53 +- 16 | 53 +- 16 | 54 +- 18 | 50 +- 17 | 0.85 | 49 |
| ***sex (female/male)*** | 40 / 16 | 26 / 9 | 6 / 4 | 7 / 3 | 0.76 | 1 / 0 |
| ***disease duration (months)*** | 5.5 +- 3.1 | 5.3 +- 3.0 | 5.05 +- 3.1 | 6.33 +- 3.9 | 0.64 | 8 |
| ***ESR (mm/hr)*** | 48 +- 30 | 55 +- 30 | 38 +- 25 | 33 +- 32 | 0.062 | 29 |
| ***CRP (ug/mL)*** | 26 +- 34 | 30 +- 28 | 20 +- 31 | 18 +- 51 | 0.53 | 0 |
| ***CCP (U/mL)*** | 341 +- 181 | 336 +- 175 | 263 +- 170 | 408 +- 189 | 0.2 | 600 |
| ***RF positive (%)*** | 52 (93%) | 33 (94%) | 9 (90%) | 9 (90%) | 0.62 | 1 (100%) |
| ***VAS*** | 65 +- 24 | 67 +- 24 | 62 +- 26 | 56 +- 23 | 0.44 | 90 |
| ***TJC*** | 11 +- 7 | 12 +- 7 | 10 +- 6 | 9 +- 8 | 0.33 | 18 |
| ***SJC*** | 8 +- 6 | 8 +- 5 | 7 +- 5 | 6 +- 7 | 0.43 | 21 |
| ***HAQ*** | 1.51 +- 0.77 | 1.61 +- 0.81 | 1.2 +- 0.58 | 1.36 +- 0.75 | 0.31 | 2.63 |
| ***DAS28*** | 5.9 +- 1.3 | 6.2 +- 1.2 | 5.7 +- 1.1 | 5.1 +- 1.1.7 | 0.078 | 7.3 |

**Supplementary Table S3 -** Number of differentially expressed genes between pathotypes in synovial tissue and peripheral blood of **ACPA-positive** RA patients, including number of upregulated genes that belong to the RA or OA risk gene set. p-value represents overrepresentation (Fisher's exact test, Methods) of upregulated RA or OA risk genes given the total number of upregulated genes. n.s. = not significant ($p > 0.05$), L = lympho-myeloid, M = diffuse-myeloid, F = pauci-immune fibroid.

| ***Synovium*** | **All genes (N = 24523)** | **RA risk genes (N = 143)** | **OA risk genes (N = 105)** | ***Blood*** | **All genes (N = 20132)** | **RA risk genes (N = 140)** | **OA risk genes (N = 76)** |
|---|---|---|---|---|---|---|---|
| *Upregulated genes in* | | | | | | | |
| *L vs M* | 1220 | 26<br>p = 8.915e-09 | 1<br>p = 0.995 | | 6 | 0<br>n.s. | 0<br>n.s. |
| *M vs L* | 695 | 0<br>n.s. | 10<br>p = 0.000804 | | 0 | 0<br>n.s. | 0<br>n.s. |
| *L vs F* | 2307 | 40<br>p = 1.800e-10 | 9<br>p = 0.6644 | | 1 | 0<br>n.s. | 0<br>n.s. |
| *F vs L* | 2627 | 8<br>p = 0.988 | 22<br>p = 0.00153 | | 0 | 0<br>n.s. | 0<br>n.s. |
| *F vs M* | 302 | 2<br>p = 0.527 | 3<br>p = 0.139 | | 4 | 0<br>n.s. | 0<br>n.s. |
| *M vs F* | 214 | 9<br>p = 4.813e-06 | 1<br>p = 0.602 | | 0 | 0<br>n.s. | 0<br>n.s. |

**Supplementary Table S4 -** Number of differentially expressed genes between pathotypes in synovial tissue and peripheral blood of **ACPA-negative** RA patients, including number of upregulated genes that belong to the RA or OA risk gene set. p-value represents overrepresentation (Fisher's exact test, Methods) of upregulated RA or OA risk genes given the total number of upregulated genes. n.s. = not significant (p > 0.05), L = lympho-myeloid, M = diffuse-myeloid, F = pauci-immune fibroid.

| ***Synovium*** | **All genes (N = 24566)** | **RA risk genes (N = 142)** | **OA risk genes (N = 105)** | ***Blood*** | **All genes (N = 18671)** | **RA risk genes (N = 137)** | **OA risk genes (N = 74)** |
|---|---|---|---|---|---|---|---|
| *Upregulated genes in* | | | | | | | |
| *L vs M* | 109 | 1<br>p = 0.46 | 0<br>n.s. | | 6 | 1<br>p = 0.0432 | 0<br>n.s. |
| *M vs L* | 131 | 1<br>p = 0.53 | 2<br>p = 0.108 | | 26 | 1<br>p = 0.174 | 0<br>n.s. |
| *L vs F* | 2 | 0<br>n.s. | 0<br>n.s. | | 0 | 0<br>n.s. | 0<br>n.s. |
| *F vs L* | 10 | 0<br>n.s. | 0<br>n.s. | | 1 | 0<br>n.s. | 0<br>n.s. |
| *F vs M* | 13 | 0<br>n.s. | 0<br>n.s. | | 73 | 3<br>p = 0.0165 | 0<br>n.s. |
| *M vs F* | 33 | 0<br>n.s. | 0<br>n.s. | | 137 | 2<br>p = 0.2661 | 0<br>n.s. |
